# A Randomised Trial of Covid-19 Transmission in Training Facilities

**DOI:** 10.1101/2020.06.24.20138768

**Authors:** Lise M. Helsingen, Magnus Løberg, Erle Refsum, Dagrun Kyte Gjøstein, Paulina Wieszczy, Ørjan Olsvik, Frederik E. Juul, Ishita Barua, Henriette C. Jodal, Magnhild Herfindal, Yuichi Mori, Solveig Jore, Fridtjof Lund-Johansen, Atle Fretheim, Michael Bretthauer, Mette Kalager, for the TRAiN study group

## Abstract

**Background:** Closed training facilities during the Covid-19 pandemic may negatively impact people’s health and wellbeing. We investigated SARS-CoV-2 virus transmission, Covid-19 and SARS-CoV-2 antibodies attributable to training facilities.

**Methods:** We randomised members aged 18 to 64 without relevant comorbidities at five training facilities in Oslo, Norway, to access or no access to their facility. Facilities opened May 22, 2020 for the training arm, applying physical distancing (1 meter for floor exercise, 2 meters for high-intensity classes) and enhanced hand and surface hygiene. We compared SARS-CoV-2 RNA status by self-administered naso-, oropharyngeal and sputum sampling after 14 days; clinical disease through electronic patient records after 21 days; and SARS-CoV-2 antibody status by dried-blood self-sampling after one month. (ClinicalTrials.gov number NCT04406909)

**Findings:** 3,764 individuals were randomised; 1,896 in the training and 1,868 in the no-training arm. In the training arm, 81.8% trained at least once, and 38.5% trained ≥six times. Of 3,016 individuals who returned the SARS-CoV-2 RNA tests (80.5%), there was one positive test. The positive individual was randomised to training, but had not used the training facility, and the workplace was identified as transmission source. There were no outpatient visits or hospital admissions due to Covid-19 in either group. Eleven individuals in the training arm (0.8% of tested) and 27 in the no-training arm (2.4% of tested) tested positive for SARS-CoV-2 antibodies (p=0.001).

**Interpretation:** Provided good hygiene and physical distancing measures, there was no increased transmission of SARS-CoV-2 at training facilities.

**Funding:** Norwegian Research Council, grant no. 312757

## Introduction

Governments and health policy makers worldwide have been taking preventive measures against Covid-19 exceeding previous pandemics (1). Social distancing such as increased distance between individuals (minimum 1 or 2 meters) is of paramount importance to contain spread of Covid-19. Many countries have closed or restricted access to schools, stores, restaurants, and work places to achieve social distancing (2).

Whilst keeping adequate distance between individuals may involve little disturbance for daily life, closures of schools, recreational activities, and work places have potentially large consequences for education, health and wellbeing, and personal and societal economy. Thus, it is important to assess social distancing measures and gain knowledge about their impacts on society as a whole (3). Due to the uncertainty of contagiousness, immunity, morbidity and mortality of Covid-19, it is unclear how to resume activities without risking increased spread of disease.

Training and exercise is important for health and wellbeing, and training facilities are important for many individuals, and for population health. In Norway, by governmental emergency law, all training facilities were closed from March 12, 2020 (4, 5). Surveys have indicated that Norwegians have a more sedentary lifestyle and exercise less after the restrictions (6). To prevent negative effects on health the effectiveness of closing on disease spread must be weighed against impact on society, and unnecessary closings prevented.

We hypothesised that the risk of SARS-CoV-2 transmission in training facilities with good hygiene and physical distancing measures would be low, and thus safe to re-open to improve health and wellbeing. This report describes the randomised testing of re-opening training facilities with close monitoring of SARS-CoV-2 transmission and disease activity to understand the impact of training facilities closure for Covid-19.

## Methods

### Study design

All training facilities in Norway were closed from March 12, 2020. For the purpose of the trial, five training facilities in Oslo, Norway, opened their premises to participants randomised to training for the period of the trial (Supplementary Appendix). All other training facilities remained closed, and participants in the no-training control arm did not have access.

### Participants

All members of the participating training facilities age 18 years or older who were not at increased risk for severe Covid-19 per criteria by the Norwegian Institute of Public Health, were eligible for participation. The criteria for high risk are at least one of the following: age 65 years or older; cardiovascular disease including hypertension; diabetes (7).

Eligible individuals were approached by email by their training facilities and signed up for the study through a secure website at the University of Oslo. Co-morbidities were self-assessed. A direct contact telephone line and email address to the study team was established for interested individuals in case of uncertainty of their medical status and other questions. All eligible individuals were informed about the nature of the trial, and provided consent before randomisation.

### Randomisation and procedures

We randomised eligible individuals to either current practice which was no access (no-training arm) to the training facility, or to access (training arm) with mitigation measures as described by the “Norwegian guidelines for Hygiene and Social Distancing in Training Facilities during the Covid-19 Pandemic”, available at https://t-i.no/wp-content/uploads/2020/04/Bransjestandard-for-sentre.pdf.

The following measures were implemented at all facilities during the trial: Avoidance of body contact; 1 meter distance between individuals at all times; 2 meter distance for high intensity activities; provision of disinfectants at all work stations; cleaning requirements of all equipment after use by participant; regular cleaning of facilities and access control by facility employees to ensure distance measures and avoid overcrowding. Changing rooms were open, but showers and saunas remained closed. Staff was present during all opening hours. Lids on trash cans were removed. Individuals were advised to stay home if they had any Covid-19 related symptoms. No masks were required, but members were advised to avoid touching their eyes, nose and mouth. Activities for the training arm included services the gyms provide ordinarily, including floor training facilities and group classes.

### Outcomes

The primary endpoint was the proportion of SARS-CoV-2 RNA positive individuals in the two study arms after 14 days. Co-primary endpoint was hospital admission in the two arms after 21 days, and secondary endpoint was the proportion of individuals with SARS-CoV-2 antibodies in the two study arms after 30 days.

### SARS-CoV-2 RNA testing

No participants were tested for Covid-19 before entering the study or the training facilities. All participants were mailed a home-test kit including two swabs and a tube with virus transport medium for SARS-CoV-2 RNA. The tests were analysed with a real-time SARS-CoV-2 RT-PCR test (Cobas®, Roche Diagnostics Inc.) at the Department of Medical Microbiology, Oslo University Hospital. Participants were instructed to sample from the oropharynx, nose and saliva according to national guidelines after median two weeks of training access in the training arm (June 8 or 9), and deliver the test to their training facility (8). Dedicated study personnel provided onsite collection of all tests, and helped with sampling for those who did not want to self-sample on June 8 and 9, 2020. The facilities remained open for individuals in the training arm until June 15, but remained closed for the no-training arm. We also offered SARS-CoV-2 RNA testing to all training facility employees working in the period.

Transmission and contact tracing of individuals positive for SARS-CoV-2 RNA were performed by trained personnel from the Norwegian Institute of Public Health.

### Clinical endpoint assessment

On June 15, 2020 (three weeks after study start), we retrieved all admissions and outpatient contacts for all somatic diagnoses (ICD-10 coding); ICU admissions, ventilator treatment, and death for all participants from the trial area hospital databases. Norway has a public, single-payer hospital system with full coverage of data for all individuals. For individuals with diagnoses which may relate to Covid-19, we contacted physicians at the respective hospitals for details to investigate if the contact was related to Covid-19.

### SARS-CoV-2 Antibody testing

Four weeks after study start (on June 24, 2020), all study participants who had provided a SARS-CoV-2 RNA test were mailed a self-sampling kit antibody testing. The participants were asked to return the dried blood spot card by mail in a prepaid envelope by June 30, 2020. Vitas Analytical Services, Oslo, Norway, prepared each sample, and analyses were performed by the Department of Immunology, Oslo University Hospital. Measurement of IgG antibodies was performed with a multiplex flow cytometric assay known as microsphere affinity proteomics (MAP) (9). Participants with insufficient quality or quantity of the dried blood spot sample or with test results close to a predefined cut-off for positivity (“borderline” results), were asked to provide a venous serum sample for analysis on Roche’s platform for SARS-CoV2 antibodies (Elecsys® Anti-SARS-CoV-2). Details are provided in Supplementary Appendix.

### Population Data on Covid-19

From publicly available sources by the Norwegian Institute of Public Health (www.fhi.no) and the Norwegian Directorate of Health (www.helsedirektoratet.no), we retrieved data on number and rates of SARS-CoV-2 positive individuals, hospital admissions, intensive care treatment and death due to Covid-19 in the trial area during the study period.

### Statistical analysis

We assumed non-inferiority of training versus no-training with regard to SARS-CoV-2 RNA positivity and hospital admission. Based on the most recent update of Covid-19 before the start of the trial from the Norwegian Institute of Public Health (May 11, 2020), we assumed that 1% in each arm would test positive for SARS-CoV-2 RNA after two weeks. We defined the smallest meaningful difference for SARS-CoV-2 transmission to be 1% between the two arms. Thus, the non-inferiority margin would be 1% for the training arm as compared to the no-training arm. For a power of 90% with an alpha of 0.05, we planned to include at least 1,696 individuals in each arm. Further power calculations for transmission rates and hospital admission are provided in the Protocol.

The primary analytic approach of the trial follows the intention-to-treat (ITT) principle. We compared the differences in event rates for the trial endpoints between the arms by chi-square test. Due to small numbers, we did not perform significance testing for all diagnosis sub-groups (Table 2). Analyses were performed using Stata Statistical Software release 16.

The study was approved by the Regional Ethical Committee of South-East Norway and the data protection officers at participating sites. All individuals provided written informed consent before enrolment. An Independent Data Safety and Monitoring Committee (DSMB) was established to ensure adequate handling of all data and trial participants.

## Results

Individuals were approached between May 15 and May 24, 2020. Randomisation of eligible individuals took place successively between May 20 and May 25, 2020, and the training facilities were opened for individuals in the training arm on May 22, 2020. In total, 3,938 individuals signed up for the trial online and provided written consent, however 113 were ineligible and 3,825 individuals were randomised. Of these, 61 withdrew consent, and thus 3764 individuals are included in the analyses; 1,896 in the training arm and 1868 in the no-training arm (Figure 1). Participant characteristics shows that the arms were well-balanced (Table 1).

**Table 1:** Characteristics of the trial participants in the training and no-training arms, and of the employees working at the training facilities during the trial. All data in numbers (%).

|  | Total | Training arm | No-Training arm |
| --- | --- | --- | --- |
| **Trial participants** | N | N (%) | N (%) |
| **Individuals** | 3,764 | 1,896 (50.4) | 1,868 (49.6) |
| **Sex** |  |  |  |
| **Women** | 1,929 | 974 (51.4) | 955 (51.1) |
| **Men** | 1,835 | 922 (48.6) | 913 (48.9) |
| **Age at enrolment** |  |  |  |
| **18-20 years** | 91 | 46 (2.4) | 45 (2.4) |
| **21-30 years** | 1,278 | 643 (33.9) | 635 (34.0) |
| **31-40 years** | 1,113 | 564 (29.7) | 549 (29.4) |
| **41-50 years** | 709 | 366 (19.3) | 343 (18.4) |
| **51-60 years** | 478 | 236 (12.4) | 242 (12.0) |
| **61-65 years** | 95 | 41 (2.2) | 54 (2.9) |
| **Training activity <sup>1</sup>** |  |  |  |
| **0 times** |  | 345 (18.2) | 1,868 (100) |
| **1-2 times** |  | 314 (16.6) | 0 |
| **3-5 times** |  | 435 (22.9) | 0 |
| **6-10 times** |  | 464 (24.5) | 0 |
| **More than 10 times** |  | 221 (11.7) | 0 |
| **Employees** |  |  |  |
| **Individuals** | 81 |  |  |
| **Sex** |  |  |  |
| **Women** | 56 |  |  |
| **Men** | 25 |  |  |
| **Age at enrolment** |  |  |  |
| **18-20 years** | 3 |  |  |
| **21-30 years** | 19 |  |  |
| **31-40 years** | 24 |  |  |
| **41-50 years** | 20 |  |  |
| **51-60 years** | 14 |  |  |
| **61-65 years** | 0 |  |  |

<sup>1</sup> Times trained at training facility during study period (data were available from four of the five facilities)

**Table 2:**
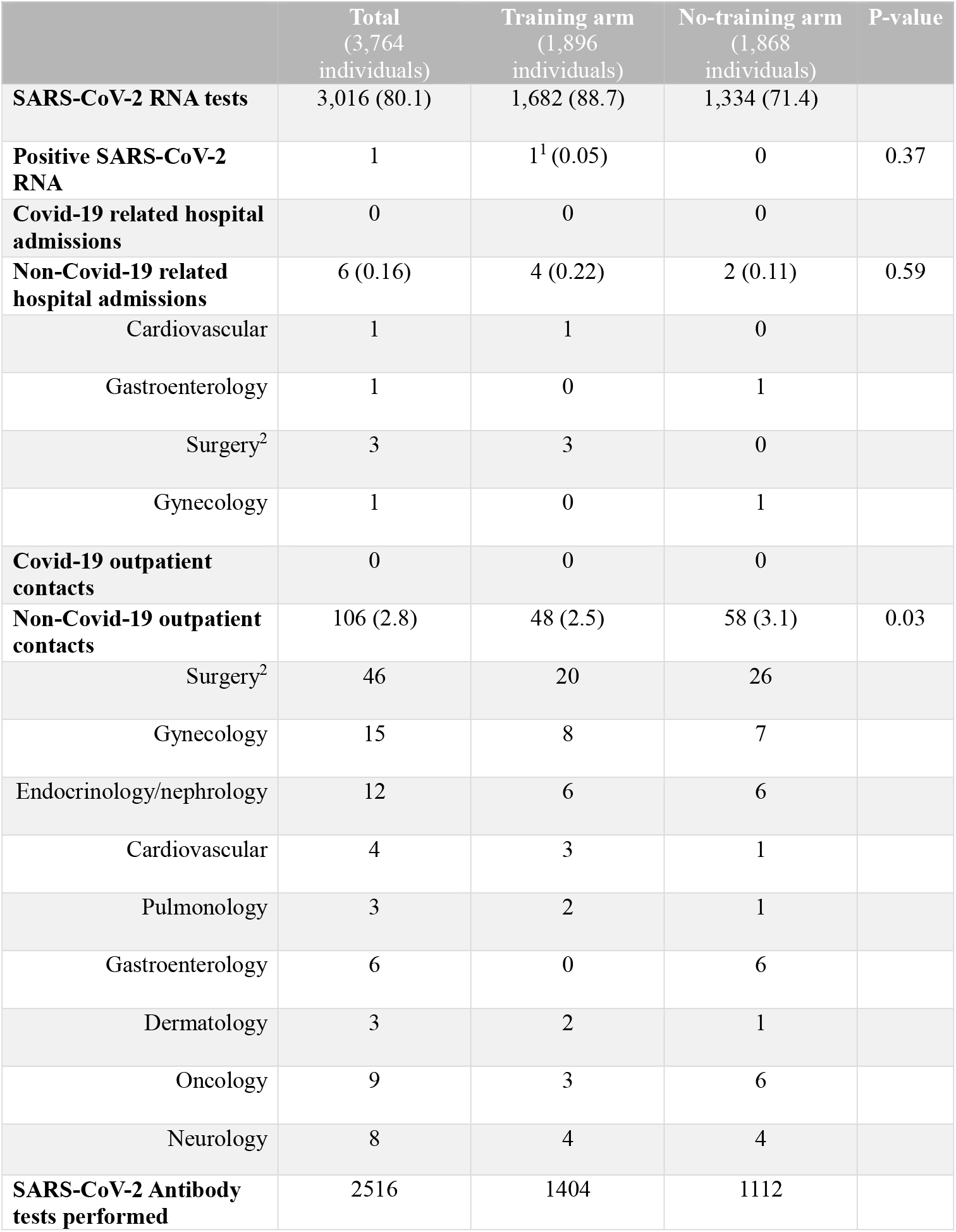
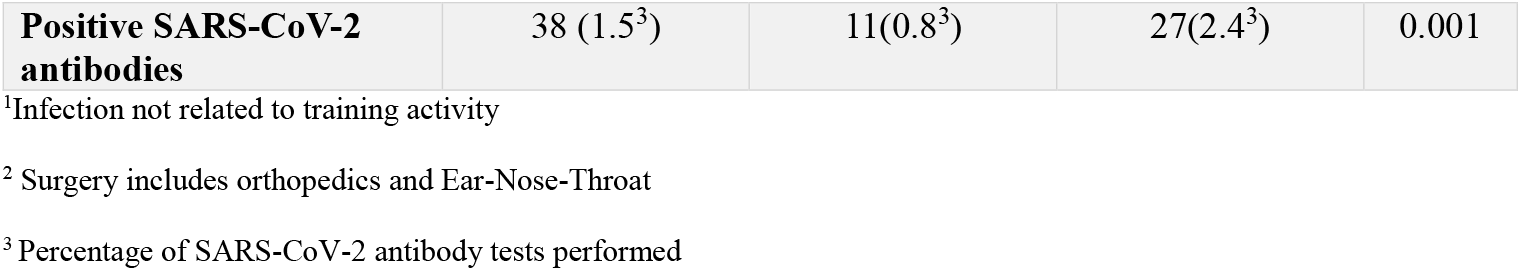
SARS-CoV-2 RNA, clinical disease and SARS-CoV-2 RNA antibodies (IgG) in the training and no-training arms. All data in numbers (%).

|  | Total<br>(3,764<br>individuals) | Training arm<br>(1,896<br>individuals) | No-training arm<br>(1,868<br>individuals) | P-value |
| --- | --- | --- | --- | --- |
| **SARS-CoV-2 RNA tests** | 3,016 (80.1) | 1,682 (88.7) | 1,334 (71.4) |  |
| **Positive SARS-CoV-2 RNA** | 1 | 1<sup>1</sup> (0.05) | 0 | 0.37 |
| **Covid-19 related hospital admissions** | 0 | 0 | 0 |  |
| **Non-Covid-19 related hospital admissions** | 6 (0.16) | 4 (0.22) | 2 (0.11) | 0.59 |
| Cardiovascular | 1 | 1 | 0 |  |
| Gastroenterology | 1 | 0 | 1 |  |
| Surgery<sup>2</sup> | 3 | 3 | 0 |  |
| Gynecology | 1 | 0 | 1 |  |
| **Covid-19 outpatient contacts** | 0 | 0 | 0 |  |
| **Non-Covid-19 outpatient contacts** | 106 (2.8) | 48 (2.5) | 58 (3.1) | 0.03 |
| Surgery<sup>2</sup> | 46 | 20 | 26 |  |
| Gynecology | 15 | 8 | 7 |  |
| Endocrinology/nephrology | 12 | 6 | 6 |  |
| Cardiovascular | 4 | 3 | 1 |  |
| Pulmonology | 3 | 2 | 1 |  |
| Gastroenterology | 6 | 0 | 6 |  |
| Dermatology | 3 | 2 | 1 |  |
| Oncology | 9 | 3 | 6 |  |
| Neurology | 8 | 4 | 4 |  |
| **SARS-CoV-2 Antibody tests performed** | 2516 | 1404 | 1112 |  |
| **Positive SARS-CoV-2 antibodies** | 38 (1.5<sup>3</sup>) | 11(0.8<sup>3</sup>) | 27(2.4<sup>3</sup>) | 0.001 |

<sup>1</sup>Infection not related to training activity
<sup>2</sup> Surgery includes orthopedics and Ear-Nose-Throat
<sup>3</sup> Percentage of SARS-CoV-2 antibody tests performed

**Figure 1:**
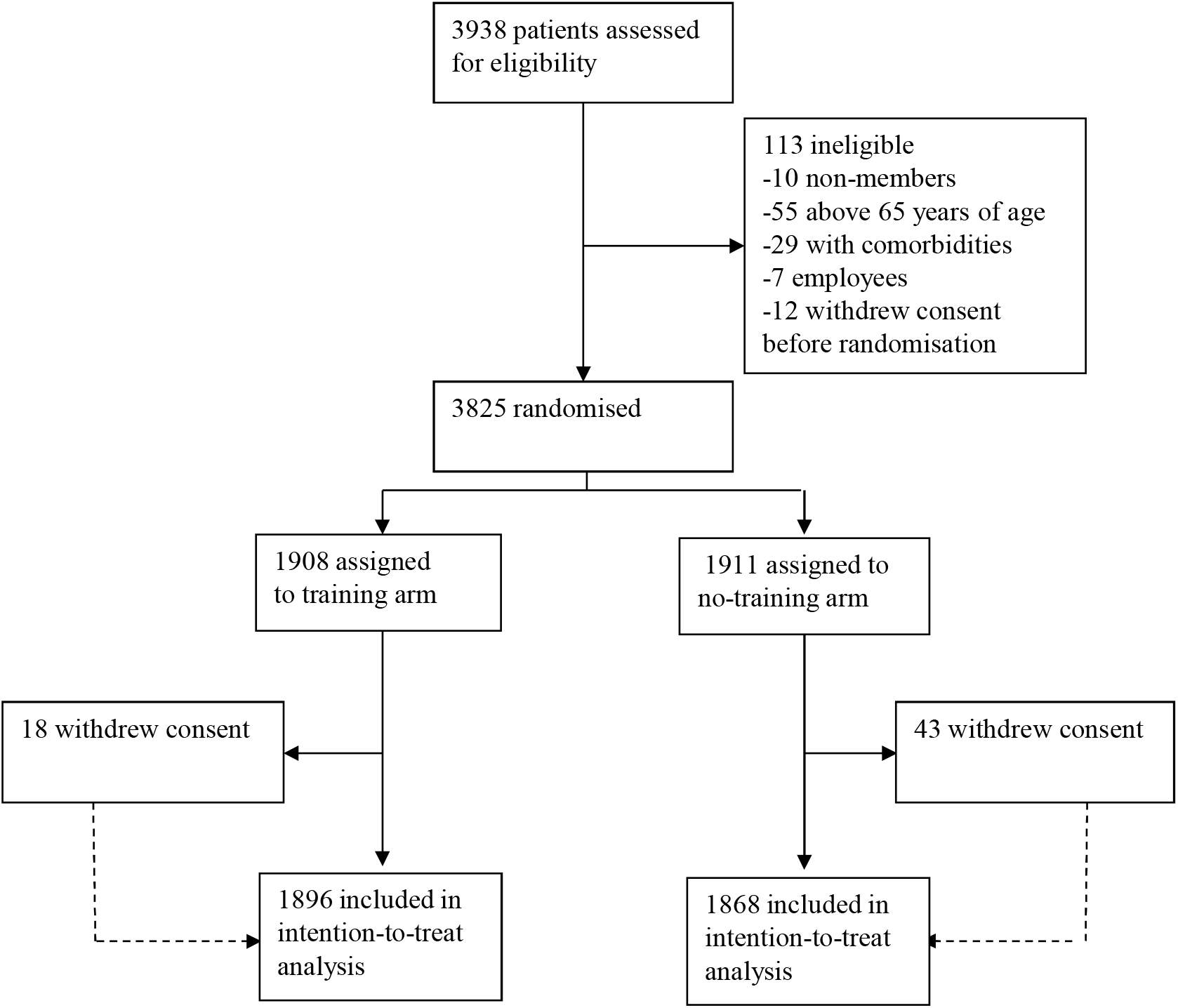
Trial profile.

### Covid-19 in Oslo during the trial

The trial area was the city of Oslo with a population of around 690 000 (10). During the first two weeks of the trial (from May 25 to June 7, 2020), 4,410 individuals in Oslo were tested for SARS-CoV-2 RNA outside the trial (11). The number of new cases was 105; 24 in the first week and 81 in the second week of the trial (rates per 100,000: 3.5 in first week, 11.7 in second week) (11). The daily number of patients who were in hospital in Oslo due to Covid-19 decreased gradually during the trial period, from 35 patients on May 22 to 21 patients on June 8, 2020.

### Training activity

Among individuals randomised to training, the majority (81.8%) trained at least once at the facility, and 38.5% trained six times or more (Table 1).

### SARS-CoV-2 RNA testing

After two weeks 3,016 participants performed sampling for SARS-CoV-2 RNA (88.7% in the training arm; 71.4% in no-training arm). There was one positive test; from an individual randomised to the training arm (Table 2, Figure 2 and 3). Transmission and contact tracing for the case revealed that the individual did not use the training facility during the trial period until the day of the sampling, but had been present at the workplace where two other individuals had tested positive for SARS-CoV-2 RNA shortly before the participant tested positive in the trial. Thus, transmission was likely unrelated to the trial intervention, and there was no further transmission during trial intervention related to the case.

**Figure 2:**
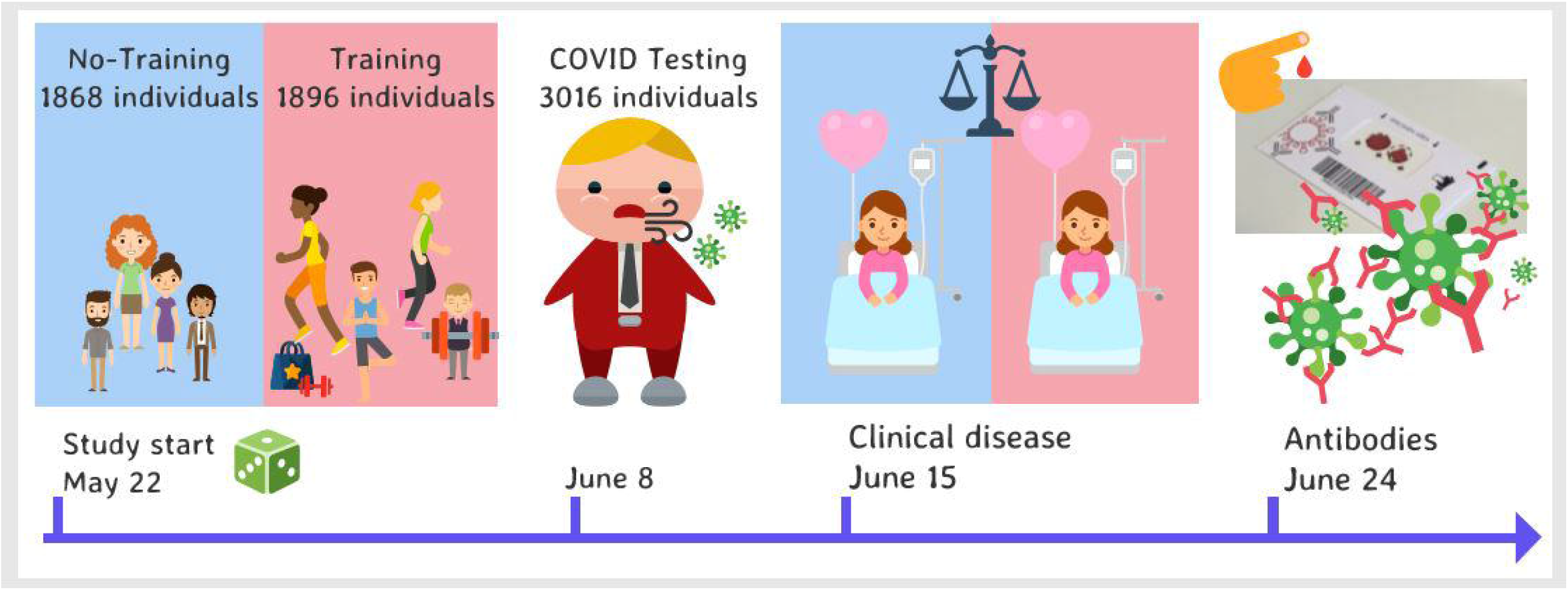
Study flowchart and graphical abstract.

**Figure 3:**
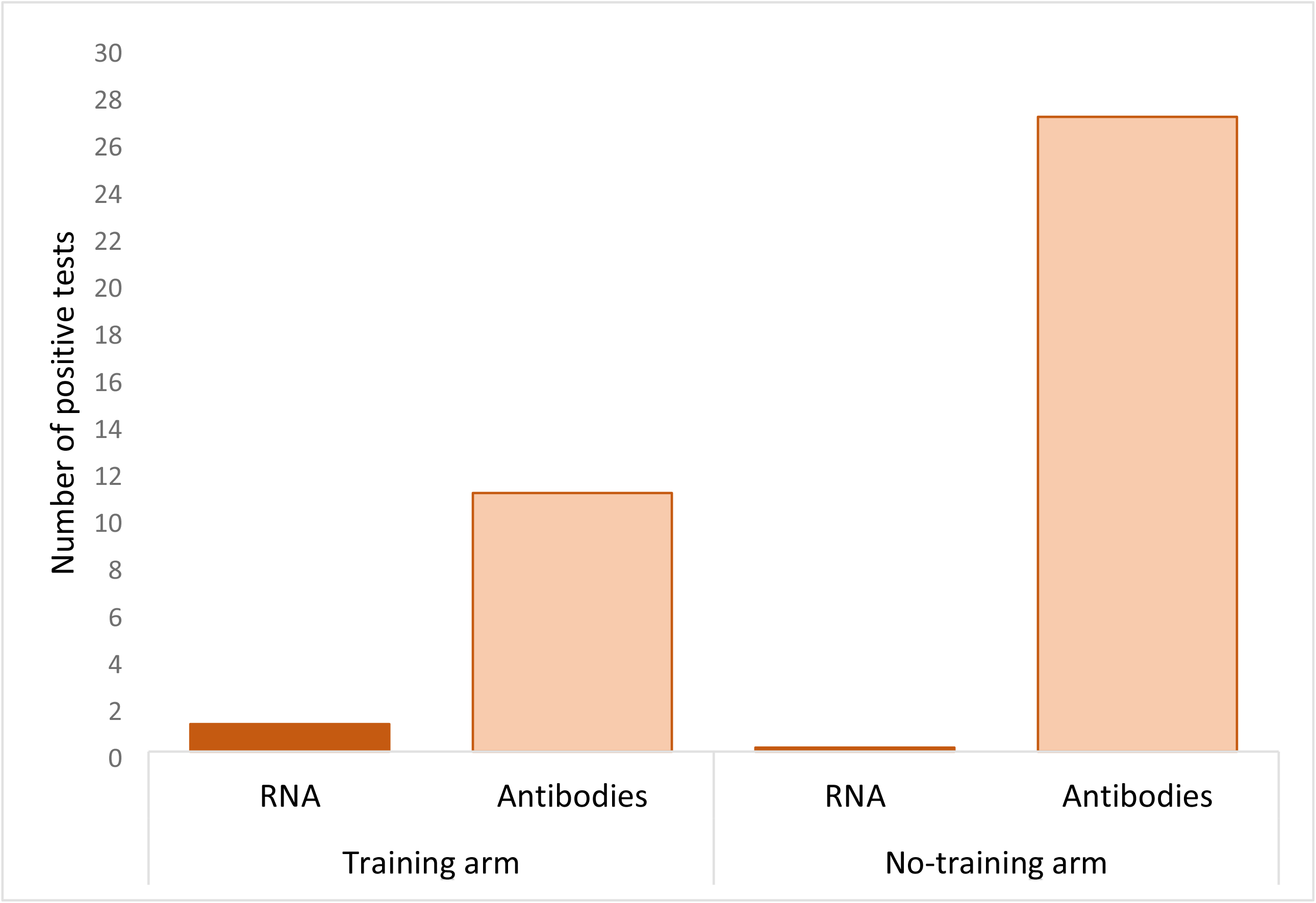
Number of positive tests for SARS-CoV-2 RNA and antibodies (IgG) in the training and no-training arm.

### Covid-19 and associated disease

A total of 106 outpatient contacts for somatic disease were registered for 106 (2.8%) participants in the hospitals serving the trial area (Table 2). There were six hospital admissions among participants; four in the training arm and two in the no-training arm. Five of the rhospital admissions were unrelated to any Covid-19 associated condition. One patient was admitted with pulmonary embolism. We contacted the attending physician who after chart review ruled out that the condition was related to Covid-19. Thus, no trial participants had hospital admissions or outpatient visits related to Covid-19 (Table 2).

### SARS-CoV-2 Antibody testing

Kits for SARS-CoV-2 antibody testing were mailed to all participants who had provided a SARS-CoV-2 RNA test; 1,682 in the training arm and 1,334 in the no-training arm. The return rate was 83.4% in both the training arm (1,404 individuals) and the no-training arm (1,112 individuals). In total, 11 individuals in the training arm (0.8% of those tested) and 27 in the no-training arm (2.4% of those tested) tested positive for SARS-CoV-2 antibodies (p=0.001) (Figure 2 and 3). Details about the antibody testing are provided in the Supplementary Appendix, Figure S1.

### Training facility employee assessment

Out of 81 employees who worked at the training facilities during the trial period and agreed to provide data, 76 (93.3%) were tested for SARS-CoV-2 RNA. None were positive. 58 employees provided antibody tests and none were positive.

## Discussion

Our trial showed no virus transmission or increase in Covid-19 or associated disease, and no increase in SARS-CoV-2 antibodies related to opening of training facilities with hygiene and physical distancing measures. The difference in SARS-CoV-2 RNA test positivity between the training and no-training arms was 0.05% (one *versus* zero cases), well below the predefined non-inferiority margin of 1%.

During the Covid-19 pandemic, many countries introduced closure of important societal activities to contain virus transmission, and by emergency law, all training facilities were closed in Norway. However, if virus containment, including contact tracing and quarantine, hand hygiene and personal physical distancing measures are sufficient to prevent virus spread, closures could be avoided. Our trial sought to test if closure of training facilities was needed. If hygienic and distancing measures could be achieved, we assumed it would be safe to open training facilities.

For the purpose of the trial, the research group and the Norwegian training facility association (Virke Training) established national mitigation guidelines for hygiene and physical distancing for training facilities in Norway in collaboration with the Norwegian Institute of Public Health. The guidelines were used in the trial and enforced by employees at the facilities at all times. The results of our trial show that with these easy and simple-to-adhere mitigations, training facilities may be allowed to re-open.

The primary concern with Covid-19 is serious disease, measured as hospital admission, need for ventilator support, and death. As a surrogate, positivity for SARS-CoV-2 RNA is often used. However, high SARS-CoV-2 RNA test positivity in individuals or groups of individuals is not necessarily a surrogate for severity of Covid-19 in a population, because SARS-CoV-2 infected individuals who do not become seriously ill or who do not transmit the disease to others who become seriously ill, may contribute to achieve immunity in the population and thus contain the disease. Therefore, we measured both SARS-CoV-2 RNA positivity and incidence of serious Covid-19 or associated disease to understand the relationship of the surrogate outcome with the clinically significant disease outcomes. As our results show, there was no increase in Covid-19-related disease due to the opening of training facilities.

Our trial was limited by the low number of events in both arms. Only one individual tested positive for SARS-CoV-2 RNA, and there was no serious Covid-19 among participants during the trial. As shown, there was indeed Covid-19 activity in Oslo during the study period, with both new cases and patients outside of the trial admitted to hospital. Our results may reflect the low risk of transmission and serious Covid-19 in healthy individuals without Covid-19 symptoms or risk factors, who were those who participated in the trial. We believe our trial population is representative of many users of training facilities and the results may thus be applied to other regions and countries (12). Although it is unclear if our findings apply to areas with much higher Covid-19 incidence rates, the rate of new positive tests outside the trial in Oslo during the trial period was not substantially different to that many states and counties in the United States reported in the same period (e.g. positive test rate per 100,000 individuals in the week of June 15 to 21, 2020 was 13 in Maine, 25 in New Jersey, and 22.5 in Massachusetts) (12).

Our sample size was based on estimates from prevalence testing in the community for Covid-19 activity. Most individuals in community testing had clinical signs or symptoms indicative of Covid-19. Thus, in accordance with evidence from population sampling in Iceland (13), we assumed considerably higher SARS-CoV-2 RNA rates in our sampling of individuals with no symptoms. This was not confirmed, as our observed rate in the trial was similar to that in the community.

Compliance with SARS-CoV-2 RNA testing was higher in the training arm (89%) than in the no-training arm (71%). However, both compliance rates are high and we consider them satisfactory to provide valid results. Disease endpoints in the trial were gathered through complete hospital registries and are not prone to self-reporting bias. Finally, the number of individuals who withdrew consent after randomisation was small (18 in the training arm and 43 in the no-training arm).

We did not observe more individuals with SARS-CoV-2 antibodies amongst individuals in the training arm, supporting the results of the RNA testing and the clinical disease data. In fact, there were significantly more individuals with detected SARS-CoV-2 antibodies in the no-training arm than in the training arm. The trial was not designed to establish any protective effect of training against Covid-19 and these results should be interpreted with caution. A possible explanation for the difference may be alternative exercise patterns in uncontrolled environments in the individuals in the no-training arm who did not have access to a training facility. This deserves further study.

It is important to perform randomised implementation and de-implementation of societal measures with large potential harms and burden for individuals and the population. Our study show that it is feasible to apply rigorous randomised testing of public health measures during an ongoing disease outbreak. The Norwegian government indeed allowed re-opening of all facilities as of June 15, 2020, provided the hygienic and social distancing measures applied in the trial can be followed. We are currently planning new randomised testing towards normal activity at Norwegian training facilities, according to the principles of rapid-cycle randomised implementation for health care services (14, 15).

## Data Availability

Data are available on request in tabulated form.

## Contributors

Conception and design: M. Bretthauer, M. Kalager, M. Løberg, L. Helsingen

Analysis and interpretation of the data: all authors

First draft of the article: M. Bretthauer

Critical revision of the article for important intellectual content: all authors

Final approval of the article: all authors

Collection and assembly of data: all authors

## Acknowledgments

The study was funded through a grant from the Norwegian Research Council, grant no. 312757. In addition to the authors mentioned above, the TRAiN study group comprised the following individuals: Anita Aalby, Madeleine Berli, Siv Furholm, Anne-Lise Horvli (all University of Oslo, Oslo, Norway); Line Norum (STOLT Training, Oslo, Norway); Halvor Lauvstad, Judit Somogyi (both EVO Training, Oslo, Norway); Alexander Myers, Tonje Poulsson, Wenche Evertsen (all SATS Training, Oslo, Norway); Hilde Sandvoll and Kjersti Oppen (both Virke Training, Oslo, Norway). The DSMB consisted of Katja Fall MD, PhD, Örebro University, Sweden, and Dag Berild MD, PHD, Oslo University Hospital. We thank the Department of Medical Microbiology and the Department of Immunology at Oslo University Hospital for virus and antibody testing, the hospitals in the Oslo region for their support with the clinical data, and all employees at the participating training facilities for their engagement and enthusiasm to establish this trial on very short notice.

### Panel: Research in context

#### Evidence before this study

We searched MEDLINE and clinicaltrials.gov on May 15, 2020, with no language restrictions, using a combination of MeSH terms and key words for Covid-19 and training facilities. We looked for clinical trials evaluating transmission of SARS-CoV-2 in training facilities, but found none.

#### Added value of this study

This is the first randomised trial testing re-opening of training facilities to understand the impact of training facilities closure for Covid-19. We found no virus transmission or increase in Covid-19 related to re-opening of training facilities with hygiene and physical distancing measures in Oslo, Norway, in May/June 2020.

#### Implications of all the available evidence

Our study show that it is feasible to apply rigorous randomised testing of public health measures during an ongoing disease outbreak. To avoid unnecessary harms and burdens for the society and the population, it is important to perform randomised implementation and de-implementation of infection preventive measures for Covid-19.

